# Complement C3 identified as a unique Risk Factor for Disease Severity among Young COVID-19 Patients in Wuhan

**DOI:** 10.1101/2020.07.24.20161414

**Authors:** Weiting Cheng, Roman Hornung, Kai Xu, Jian Li

## Abstract

**Background:** Given that a substantial proportion of the subgroup of COVID-19 patients that face a severe disease course are younger than 60 years, it is critical to understand the disease-specific characteristics of young COVID-19 patients. Risk factors for a severe disease course for young COVID-19 patients and possibly non-linear influences remain unknown.

**Methods:** Data of COVID-19 patients with clinical outcome in a designated hospital in Wuhan, China, collected retrospectively from Jan 24^th^ to Mar 27^th^, were analyzed. Clinical, demographic, treatment and laboratory data were collected from patients’ medical records. Uni- and multivariable analysis using logistic regression and random forest, with the latter allowing the study of non-linear influences, were performed to investigate and exploit the clinical characteristics of a severe disease course.

**Results:** A total of 762 young patients (median age 47 years, interquartile ranges [IQR] 38 - 55, range 16 - 60; 55.9% female) were included, as well as 714 elderly patients as a comparison group. Among the young patients, 362 (47.5%) had a severe/critical disease course and the mean age was significantly higher in the severe subgroup than in the mild subgroup (59.3 vs. 56.0, Student’s t-test: p < 0.001). The uni- and multivariable analysis suggested that several covariates such as elevated levels of ASS, CRP and LDH, and decreased lymphocyte counts are influential on disease severity independent of age. Elevated levels of complement C3 (odds ratio [OR] 15.6, 95% CI 2.41-122.3; p=0.039) are particularly associated with the risk for the development of severity specifically in young patients, where no such influence seems to exist for elderly patients. Additional analysis suggests that the influence of complement C3 in young patients is independent of age, gender, and comorbidities. Variable importance values and partial dependence plots obtained using random forests delivered additional insights, in particular indicating non-linear influences of risk factors on disease severity.

**Conclusion:** In young patients with COVID-19, the levels of complement C3 correlated with disease severity and tended to be a good predictor of adverse outcome.

## Introduction

The pandemic caused by the coronavirus disease 2019 (COVID-19) that is associated with the severe acute respiratory syndrome coronavirus 2 (SARS-CoV-2) has affected almost every corner of the world. As of June 30^th^, 2020, there have been more than 10 million confirmed cases, including more than 500,000 deaths according to a report by the World Health Organization (WHO)^1^, where the numbers of cases and deaths are expected to continue to rise. The clinical spectrum of COVID-19 appears to be wide, encompassing asymptomatic infection, mild upper respiratory illness, neurological symptoms, renal and gastrointestinal complications, severe viral pneumonia with respiratory failure, multiple organ failure and even death^2-5^. Approximately 20∼25% of patients will have a severe disease course.

Despite numerous studies showing a higher risk of severe COVID-19 in elderly patients, a substantial proportion of young patients also have an increased risk of developing a severe course. According to a report of U.S. CDC, 47% of hospitalized patients are under the age of 65, and 48% of those admitted to ICU are under the age of 65^6^. Although potential risk factors for mortality were reported to include advanced age, male gender, presence of comorbidities, the development of a cytokine storm and an immunocompromised status^8-9^, the risk factors for the development of a severe course specific to young patients (<= 60 years old) remain under investigation. Of note, to date, a large proportion of studies applied logistic regression to analyze risk factors related to COVID-19 infection, assuming an underlying causal linear influence on the log odds^2-5^. However, given the intricate complexity of COVID-19 infection, statistical analysis with the consideration of non-linear relationships might provide more insightful information on COVID-19 related potential risk factors.

In an effort to fill these gaps, an important aim of this single-center study was to analyze clinical, demographic and treatment data of patients sequentially admitted into the Wuhan No.1 hospital, in an attempt to elucidate risk factors and main causes among young COVID-19 patients for experiencing a severe disease course. A further aim was to use the large amount of data available in this study to foster the knowledge on general, age-independent, and non-linear relational risk factors for a severe disease course.

## Methods

### Study design and participants

This retrospective, single-center cohort study involved adult patients who were diagnosed with COVID-19 pneumonia between Jan. 24^th^ and Mar 27^th^, 2020, in the major government designated hospital in Wuhan: Wuhan No.1 Hospital. The date of the last follow-up was April 8^th^, 2020. The primary outcome was severity at the end of the study period. All patients were residents of Wuhan, and the diagnostic criteria of COVID-19 were based on the Diagnosis and Treatment Protocol for the 2019 Novel Coronavirus Pneumonia published by the National Health Commission of China.

The newly diagnosed patients were required to meet one of the following conditions: (1) positive signals of COVID-19 nucleic acids were found in the detection of fluorescent real-time RT-PCR; (2) the detection of viral gene sequencing showed a high degree of homology with the new coronavirus COVID-19. Patients with mild symptoms were required to meet the following conditions: (1) history of epidemiology; (2) fever or other respiratory symptoms; (3) abnormalities of typical CT images of viral pneumonia. Patients with a severe condition met one of the following conditions: (1) shortness of breath, respiratory rate≥30 times/min; (2) oxygen saturation (resting state) ≤93%; (3) PaO2/FIO2≤300mm Hg. Critically ill patients were required to meet one of the following conditions: (1) respiratory failure, mechanical ventilation required; (2) shock; (3) combined with other organ failure, and ICU monitoring treatment required.

The following data were collected on admission: age, sex, symptoms from onset to hospital admission (fever, cough, dyspnea, myalgia, rhinorrhea, arthralgia, chest pain, headache, and vomiting), comorbidities (cardiovascular disease, chronic pulmonary disease, cerebrovascular disease and chronic neurological disorders, diabetes, malignancy, and smoking), vital signs (heart rate, respiratory rate, and blood pressure), laboratory values on admission (serum hemoglobin concentration, lymphocyte counts, platelet counts, diverse protein markers), treatment regime used for COVID-19 pneumonia (antiviral agents, antibacterial agents, and Chinese medicine), date of symptom onset, admission, virus testing, CT-scan, as well as condition improvement and living status. The study was approved by the Ethics Committee of Wuhan No.1 Hospital (No. 202008).

### Treatment Protocol for SARS-CoV-2 Pneumonia

The treatment strategy for patients with COVID-19 pneumonia was based on the guidelines of the world health organization (WHO)^10^, which included symptom relief, treatment of underlying diseases, prevention of superimposed bacterial infections, active prevention of complications such as sepsis and ARDS and support organ vital function in a timely fashion. Oxygen supplementation was provided for patients with desaturation by means of high flow oxygen via nasal prong, non-invasive and invasive mechanical ventilation, or extracorporeal membrane oxygenation (ECMO) if required.

### Statistical Considerations

The outcome in this study was whether or not the patients experienced a severe to critical course of the disease. This outcome will be denoted “severe vs. mild” in the following. All eligible variables were considered as covariates potentially influencing the outcome in the statistical analysis (supplement section “In-depth description of the statistical analysis flow”). Univariable analysis was performed using logistic regression analysis, where p-values were adjusted for multiple testing by means of the Benjamini-Hochberg procedure. The popular multiple imputation approach MICE^11^ was employed to deal with missing values in the multivariable analyses, where 20 imputed data sets were used in each analysis (m = 20). The ratio of CRP versus ALB and CRP, as well as WBC and ANC correlated very strongly (*ρ* > 0.9), which is why CRP and ANC were not considered in the multivariable analysis. The latter analysis was performed separately for the young patients and for all patients together 1) using logistic regression in combination with an automatic forward covariate selection procedure based on the Akaike information criterion (AIC)^12^ applicable to multiply imputed data^13^ and 2) random forest^14^. As a sensitivity analysis^15^, the Bayesian information criterion (BIC)^16^ that tends to select fewer covariates than the AIC was also considered and backward selection was performed in addition, both, when using the AIC and the BIC. The prediction performance of the models was estimated using 20 times repeated stratified K-fold cross-validation (K = 3,4,5), repeating the whole model selection process on each training set in each cross-validation iteration, excluding the corresponding test set^17^. Multiple imputation was performed separately on training and test sets^18^. As prediction performance measures the AUC and the Brier score were used. Random forest was also used to rank the covariates with respect to their importance for prognosis via the AUC covariate importance values^19^ and to estimate the influence forms of the covariates using partial dependence plots (PDPs)^20^. All statistical analyses were performed using the software R, version 3.6.3. All p-values smaller than 0.05 were considered as statistically significant. For further details and explanations, the interested reader is referred to the detailed description of the statistical analysis flow in the supplement (section “In-depth description of the statistical analysis flow”).

## Results

### Demographic and Clinical Features

The young cohort (age <= 60 years old) consisted of 762 patients (median age 47 years, interquartile ranges [IQR] 38 - 55, range 16 - 60; 55.9% female), who were admitted between Jan. 2020 and Mar. 2020 to Wuhan No.1 Hospitals. As shown in Table 1, 400 (52.5%) of the young patients had a mild condition during hospitalization (mild subgroup), while 362 (47.5%) developed a severe or critical disease course (severe subgroup). The mean age was significantly higher in the severe subgroup than in the mild subgroup (59.3 vs. 56.0, Student’s t-test: p < 0.001). 145 (19.8%) patients were affected by underlying diseases, hypertension (14.3%) being the most common one (Table 1). The median body temperature on admission was 36.6°C (IQR 35.6 - 37.2), no difference of median temperature between both subgroups (mild vs. severe) was seen. The hospitalization in the severe subgroup was significantly longer than that in the mild subgroup (20.0 days [IQR 15.0 - 25.0] vs. 8.0 days [IQR 5.0 - 13.0]). The majority of patients received Chinese traditional medicine (93.4%), antiviral treatments (86.7%), and antibiotics (75.3%). In this cohort, the most frequently applied oxygen therapy was UOC (78.2%). All key laboratory findings of this cohort are listed in Table 1. Additionally, an elderly cohort of patients (median age 69 years [IQR] 65 - 75, range 61 - 97, 52.8% female) was included, mainly for statistical comparisons with the young cohort.

**Table 1:**
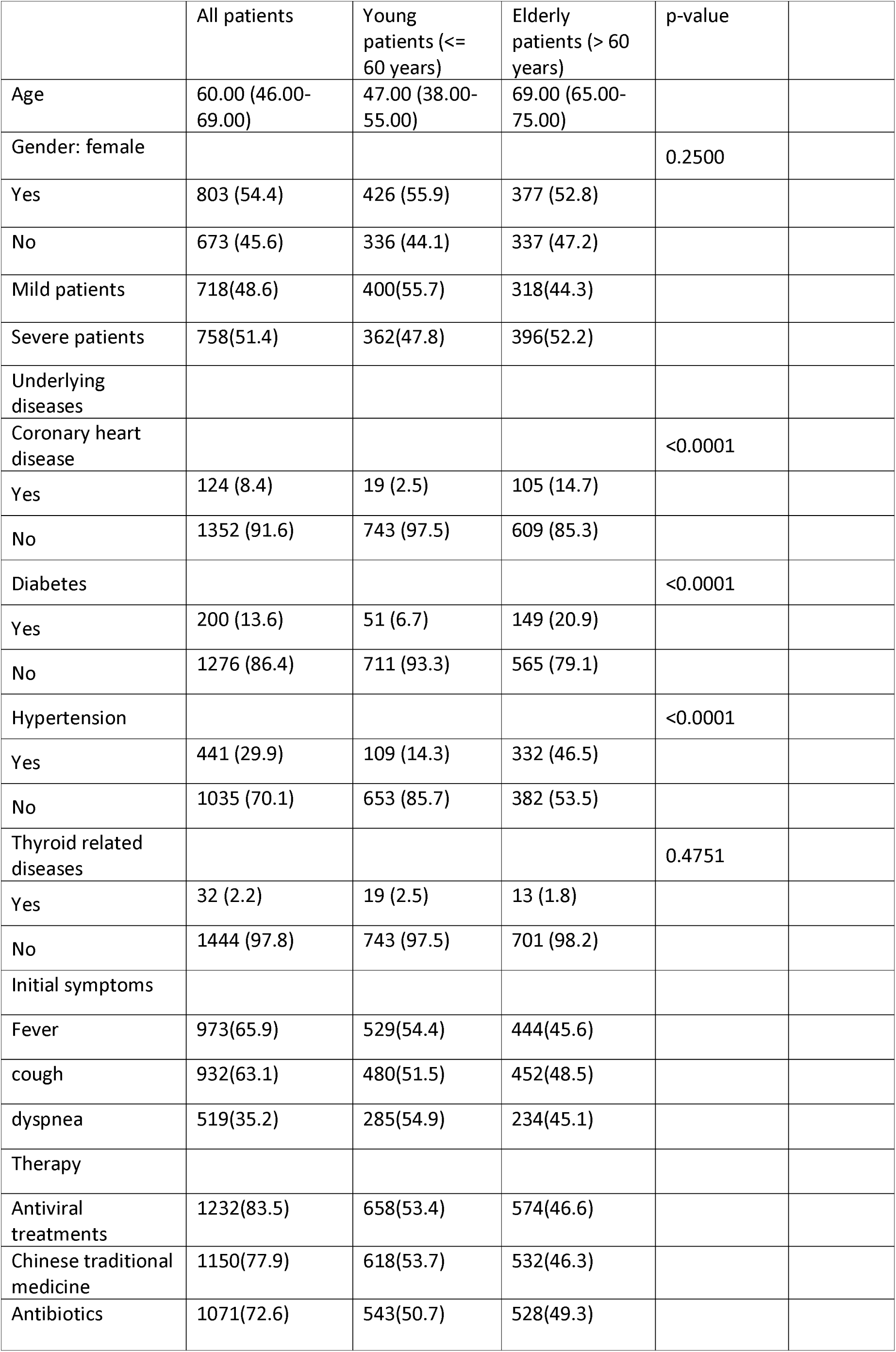
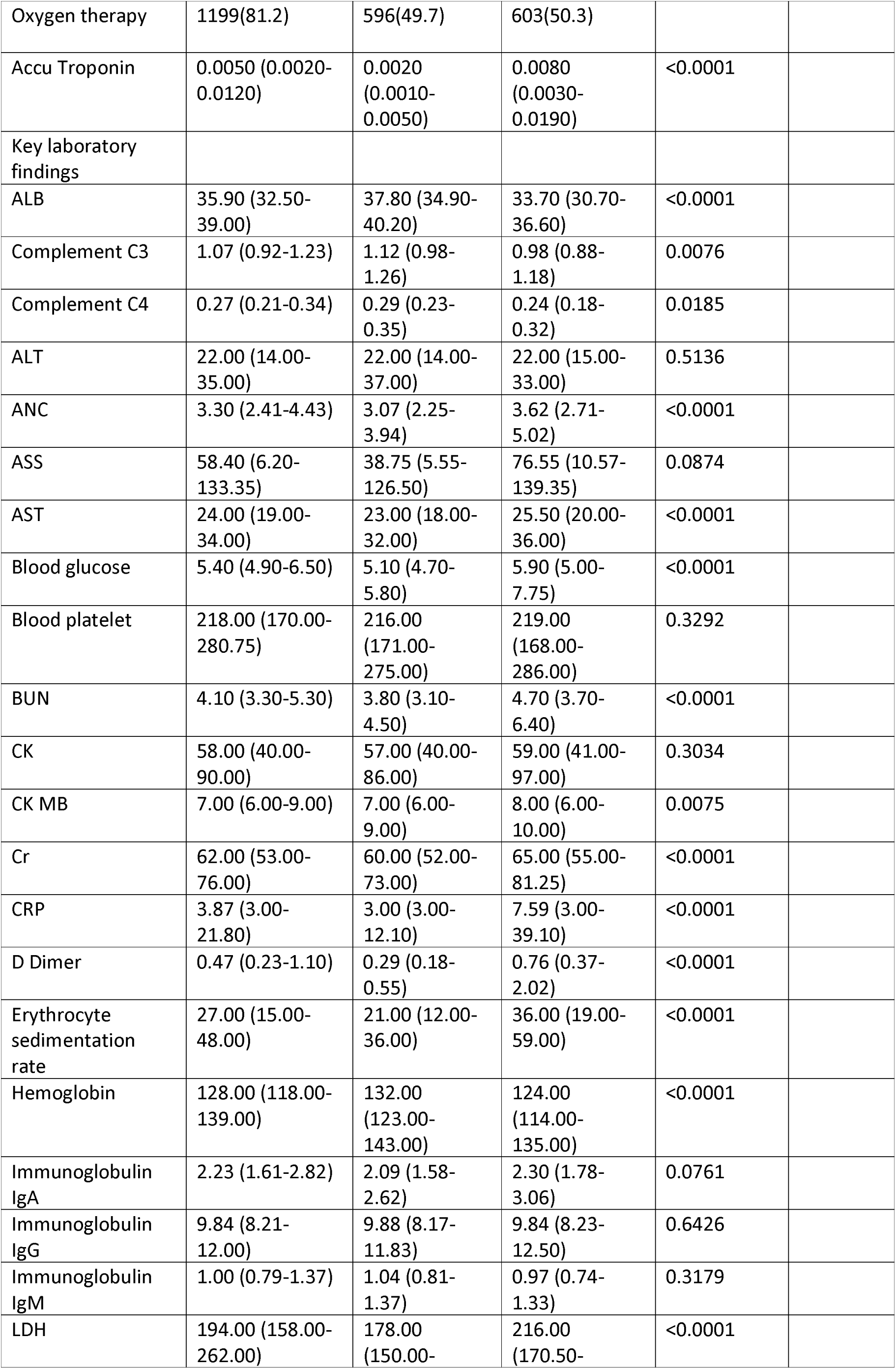
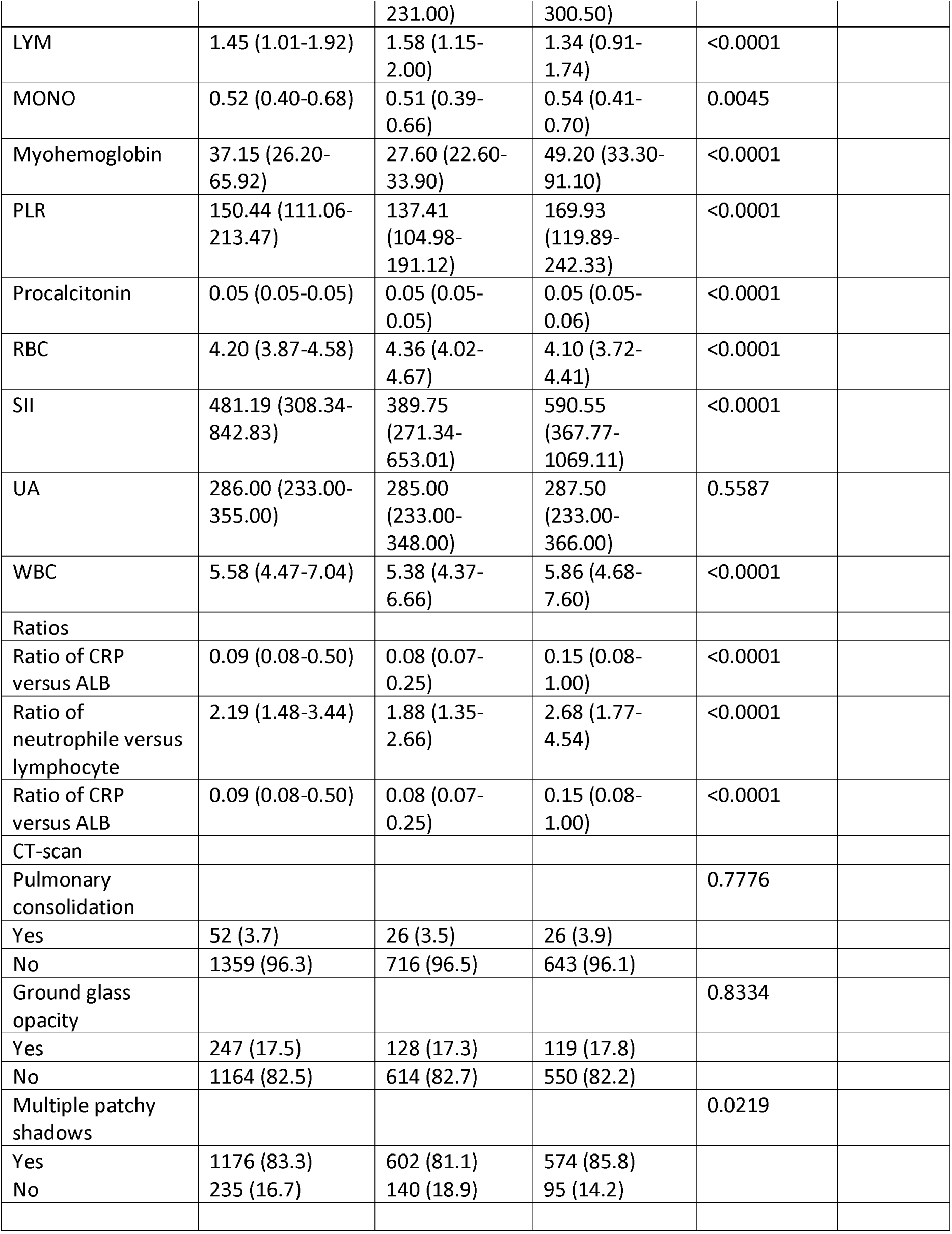
Clinical and demographic characteristics, treatment and key laboratory findings. Metric covariates are reported as medians with interquartile ranges in the following form: ‘Median (First Quartile-Third Quartile)’. Categorical covariates are reported as percentages in the following form: ‘Absolute number (Percentage)’. Differences between the young and elderly patients were tested using the Wilcoxon test and Fisher’s exact test for metric covariates and categorical covariates, respectively.

### Potential Risk Factors Associated with Disease Severity in the Young Patients

In the univariable analysis, elevated level of complement C3, SII, CRP, ASS, LDH, the ratio of CRP versus ALB, and the ratio of neutrophile versus lymphocyte were significantly associated with an increased risk for the development of a severe disease course of COVID-19 infection in the young patients (Table 2; Supplementary Table 1 contains the non-significant results in addition). In contrast, decreased levels of ALB and LYM were significantly correlated with the development of a severe disease course. ASS had the largest covariate importance value, both for young and elderly patients. The covariate importance values (Figure 1) and the PDPs (Figure 2, Supplementary Figures 1 to 4) obtained using random forest analysis suggest that complement C3 is only prognostic in young patients.

**Table 2:**
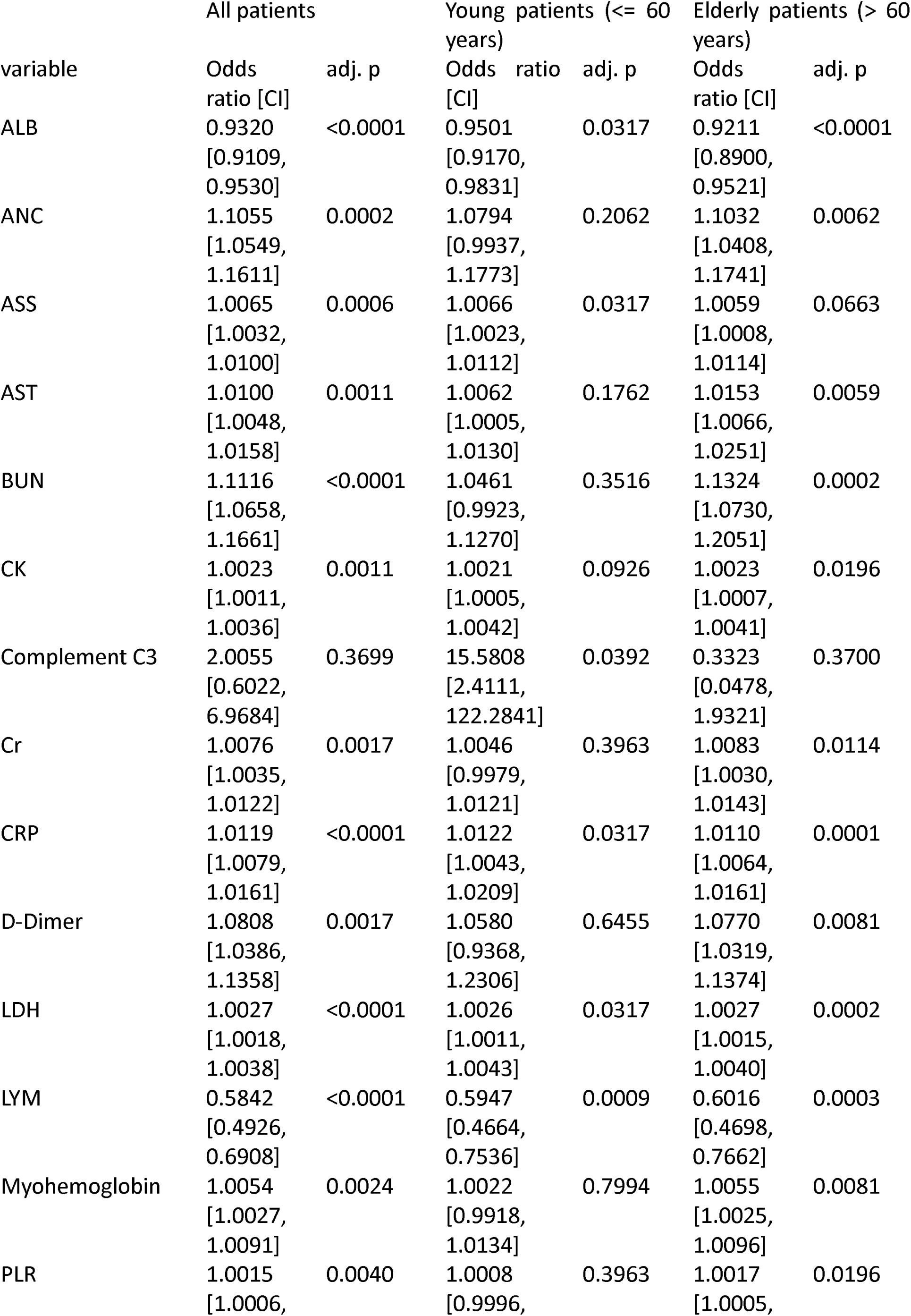
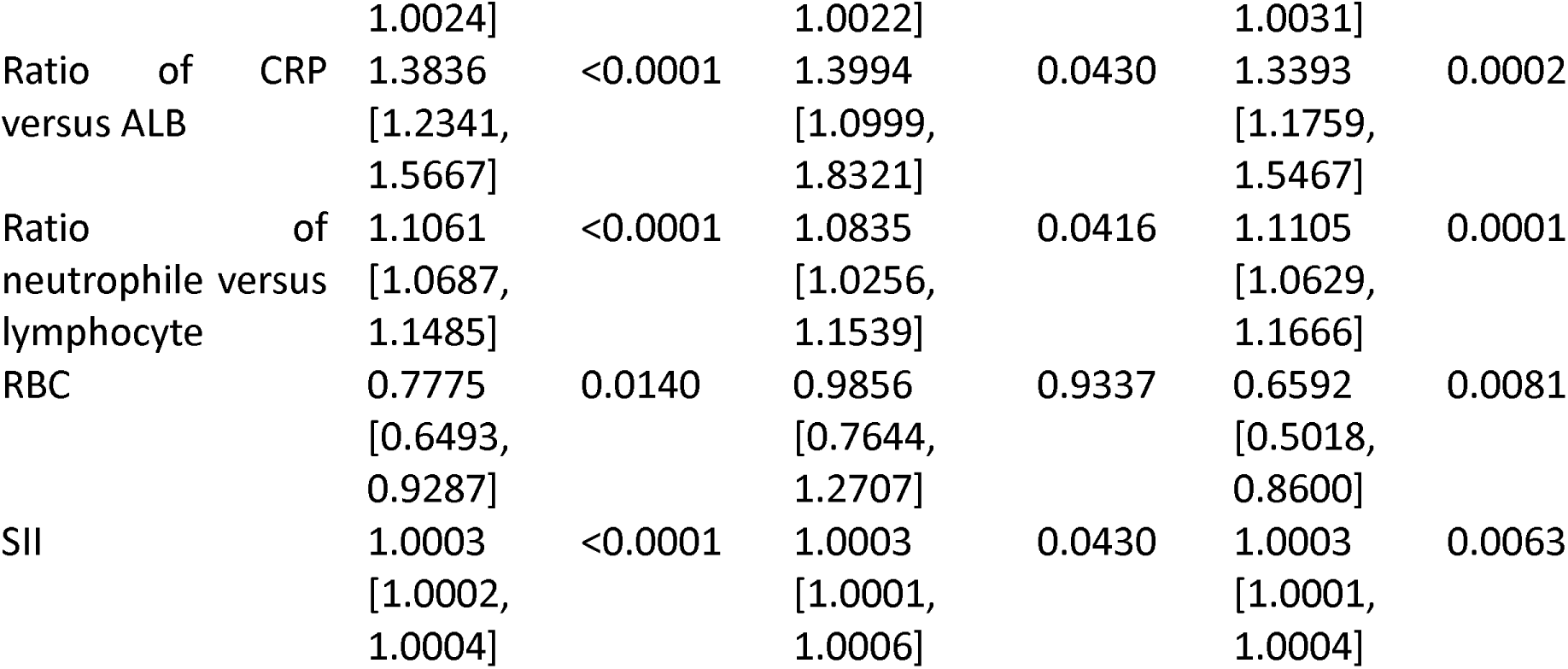
Univariable logistic regression for the outcome “severe vs. mild”. The p-values were adjusted for multiple testing separately for the analysis of all patients, young patients, and elderly patients. Increased levels of complement C3 and ASS were associated with an increased risk for severity only in the young patient cohort. Supplementary Table 1 contains the results for all covariates.

**Figure 1:**
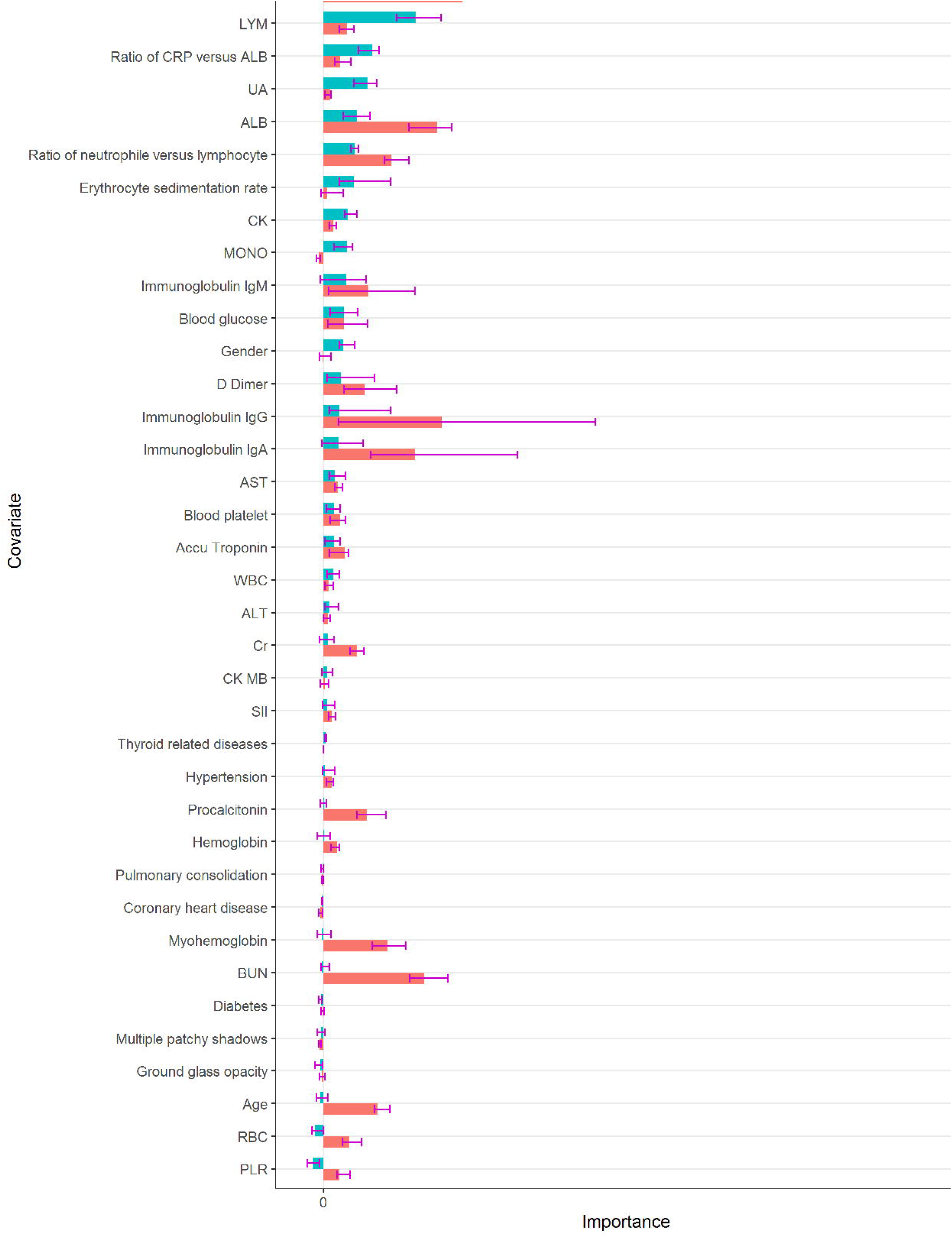
AUC variable importance with respect to predicting the outcome “severe vs. mild” for young and elderly patients calculated using random forests. The larger the importance value of a covariate is, the greater the improvement of prediction performance by including this covariate in prognosis tends to be. Complement C3, C4, LYM, the ratio of CRP versus ALB, and UA seem to influence the prognosis of the development of severity mostly in young patients. The bars show the medians of the 20 importance values calculated using the 20 imputed data sets from the multiple imputation. The error bars illustrate the variabilities of the importance values: The lower / upper ends show the first / third quartiles of the 20 importance values, that is, 25% percent of the importance values lie below / above these values. To make the raw importance values comparable between young and elderly patients, both for the young and for the elderly patients, the raw importance values were divided by the means of all importance values with positive sign.

**Figure 2:**
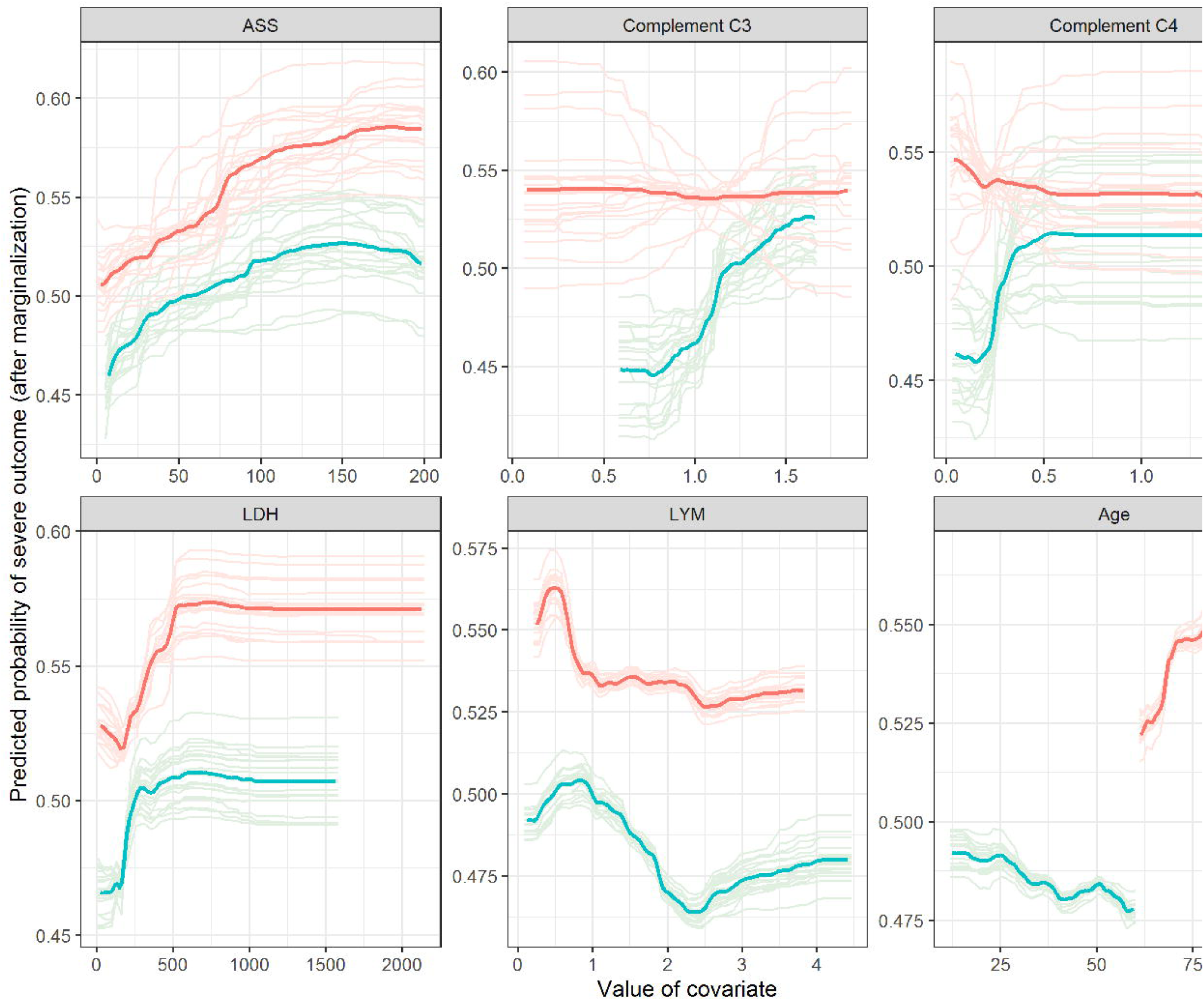
Partial dependence plots (PDPs) for young and elderly patients calculated using random forests. In simplified terms, a PDP shows the influence of a covariate on the outcome after adjusting for the influences of the other covariates. The PDPs for the five variables with largest AUC importance values in young patients and that for ‘Age’ are shown. The light lines show the 20 individual PDPs calculated using the imputed data sets from the multiple imputation. The bold lines show averages over the 20 individual PDPs.

Generally, the covariate importance values reveal that, while the number of relevant risk factors is larger for the elderly patients, the difference in importance between the most important risk factors and the remaining risk factors is more pronounced for the young patients. Supplementary Figures 5 to 10 show the corresponding variable importance values and PDPs obtained for all patients, irrespective of age. Many of the PDPs indicate complex influence forms of the covariates. While ASS again is associated with the largest importance values here, the importance values of complement C3 are relatively small. The latter confirms that a higher level of complement C3 probably is a risk factor only associated with severity in young patients. After obtaining the latter result, additional analyses were performed in order to investigate, whether the influence of complement C3 is different for specific subgroups of young patients (see supplement section “Subgroup analysis of the influence of complement C3 in young patients” for details). The results of these analyses did not suggest any relevant dependence of the influence of complement C3 in young patients on age, gender, and comorbidities, indicating that this risk factor is relevant for young patients independent of their specific characteristics (Supplement Figures 11-13).

### Multivariable Statistical Model-Analysis for Disease Severity

The results of the multivariable analysis using logistic regression in combination with the forward selection algorithm and the AIC criterion showed that the risk for the development of a severe disease course for COVID-19 in young patients was higher for combinations of elevated levels of complement C3, increase of ASS and SII, and reduced levels of LYM, PLR and UA (Table 3). Applying the backward selection algorithm, UA was not selected in the multivariable model due to lack of importance, but gender, hypertension, thyroid related disease, and BUN were selected instead, indicating potential importance to the risk for development of a severe disease course in the young patients (Supplementary Table 2). However, a sensitivity analysis presented in the supplement (section “Model stability analysis”) indicates that the results obtained using the forward selection algorithm (Table 3) may be more statistically stable. When using the BIC criterion instead of the AIC criterion, only complement C3 and ASS were shown to be relevant to the severity in these young patients (Supplementary Table 3).

**Table 3:**
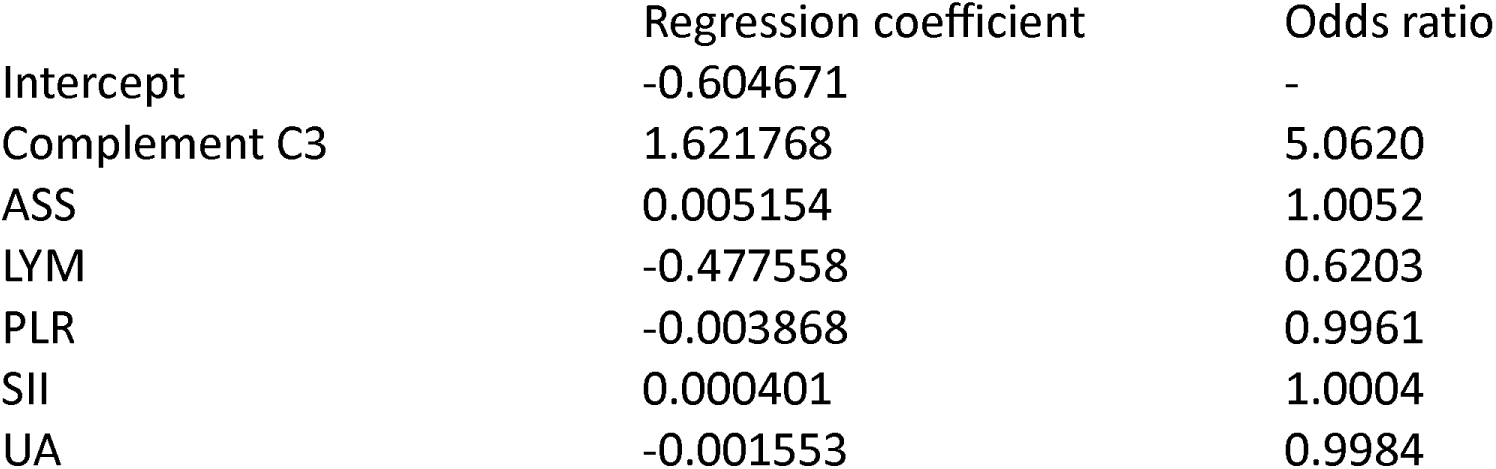
Multivariable logistic regression models for the outcome “severe vs. mild” in young patients selected using the AIC criterion and forward selection.

The results of multivariable logistic regression for all patients (including young and elderly) using the AIC criterion and the forward selection algorithm differed partly from that obtained for the young patients. The risk for the development of a severe disease course in all patients was high for increased levels of complement C3, ASS, BUN, LDH and immunoglobulin G (IgG) and decreased levels of ALB, PLR and IgA (Table 4). Note that the fact that complement C3 was included in the forward selection for all patients is very likely only due to its importance within the young cohort: In the univariable analysis complement C3 was only significant for the young patients and, as seen in Figure 1, the covariate importance value of complement C3 is only large for the young patients. As revealed by the covariate importance values obtained through the random forest analysis (Figure 1), increased levels of ASS seem to be similarly associated with the development of a severe disease course in elderly patients as well as in young patients. When using the backward selection algorithm, LYM and blood platelet were selected in addition, indicating potential relevance associated with disease severity (Supplementary Table 4). Using the BIC criterion together with the forward selection algorithm, ASS, BUN, ALB, and LDH were selected, where the corresponding odds ratios (Supplementary Table 5) were very similar to those obtained in the model obtained using the AIC (Table 4). Applying the BIC criterion together with the forward selection algorithm delivered the same result.

**Table 4:** Multivariable logistic regression models for the outcome “severe vs. mild” in all patients irrespective of age selected using the AIC criterion and forward selection.

|  | Regression coefficient | Odds ratio |
| --- | --- | --- |
| Intercept | 0.191987 | - |
| Complement C3 | 0.506456 | 1.6594 |
| Immunoglobulin IgA | -0.134694 | 0.8740 |
| Immunoglobulin IgG | 0.035108 | 1.0357 |
| ALB | -0.044892 | 0.9561 |
| ASS | 0.00574 | 1.0058 |
| BUN | 0.06745 | 1.0698 |
| LDH | 0.001668 | 1.0017 |
| PLR | -0.000795 | 0.9992 |

**Table 5:** Performances of the models measured using stratified K-fold cross-validation. For each value of K, the stratified K-fold cross-validation was repeated twenty times and the results averaged. Higher values of the AUC and smaller values of the Brier score are preferable.

|  | K in cross-validation | AUC |  |  | Brier score |  |  |
| --- | --- | --- | --- | --- | --- | --- | --- |
|  |  | Logistic regression - AIC | Logistic regression - BIC | Random forest | Logistic regression - AIC | Logistic regression - BIC | Random forest |
| Young patients | 3 | 0.5512 | 0.5542 | 0.5962 | 0.2740 | 0.2584 | 0.2438 |
|  | 4 | 0.5405 | 0.5454 | 0.5847 | 0.2764 | 0.2606 | 0.2462 |
|  | 5 | 0.5327 | 0.5344 | 0.5838 | 0.2772 | 0.2640 | 0.2461 |
| All patients | 3 | 0.6069 | 0.6186 | 0.6310 | 0.2502 | 0.2425 | 0.2369 |
|  | 4 | 0.6027 | 0.6140 | 0.6271 | 0.2525 | 0.2444 | 0.2378 |
|  | 5 | 0.6059 | 0.6156 | 0.6276 | 0.2500 | 0.2433 | 0.2378 |

In summary, the estimated prediction performances of the multivariable logistic regression models for all patients were better than those of the models obtained specifically for the young patients (Table 5). The random forest, that considers also non-linear influences, performed best and the model selected using the BIC (that included less covariates) better than that selected using the AIC. Supplementary Table 6 provides an overview on which covariates were selected in each of the models obtained using the AIC and BIC criterion with forward and backward selection.

## Discussions

COVID-19, caused by the SARS-CoV-2 virus, is a public health event that poses a serious threat to human health, and the number of COVID-19 cases in young adults is higher than expected: According to the US CDC report almost half of the patients are younger than 65, and young patients have a substantial risk for development of a severe disease course. Facing the rapidly evolving circumstances caused by the COVID-19 pandemic, it is essential to prioritize the medical resources by effectively conducting clinical stratification of COVID-19 young patients. Therefore, it is critical to early identify and investigate potential risk factors for the development of a severe/critical disease course during COVID-19 infection. In order to shed light on potential risk factors associated with a severe disease course in young patients, this study investigated clinical, demographic, treatment, and laboratory data from a group of COVID-19 patients using a set of comprehensive modern statistical methodologies. The univariable analysis suggested that potential risk factors for disease severity in young patients (age<=60 years; n=762) are in part different from that in elderly patients (age>60 years; n=714). Specifically, elevated levels of complement C3 and ASS were only significantly associated with higher risks for a severe disease course in the young patients. In contrast, increased levels of ANC, AST, BUN, CK, CR, D-dimer, Myo-hemoglobin, and PLR, and decreased levels of RBC had a significant influence on this risk only for elderly patients. Even though ASS lacked a statistical significance in the elderly subgroup, the covariate importance values (Figure 1) suggest that ASS is an important risk factor irrespective of patients’ age. While complement C4 missed significance in both age groups, the covariate importance values and PDPs suggest that this covariate, complement C3, could be a particularly strong risk factor in the young cohort, while its influence was not significant in elderly patients. Further analysis showed that the significance of complement C3 in young patients was independent of the common risk factors age, gender and the presence versus absence of comorbidies. Risk factors reported previously in COVID-19 infection such as CRP^7^, lactate dehydrogenase^3^ and decreased lymphocyte^8^ were validated both in young and elderly patient cohorts. Only the D-dimer and procalcitonin did not show significance in the young patient cohort. The PDPs confirmed the observed differences and revealed that the influences of many covariates on the log odds of disease severity of COVID-19 infection are strongly non-linear. However, in logistic regression it is assumed that these influences are linear, which is likely an important reason why the random forests outperformed the multivariable logistic regression models both for the cohort of young patients and for all patients taken together.

The complement family is an important integral component of the innate immune response to viruses, not only protects the body from infectious agents such as viruses and bacteria, but also plays a key role in promoting inflammatory processes triggering inflammatory cytokine storm^21^. The abnormal activation of various innate immune pathways, such as complement system, cytokines and thrombosis pathways, is considered as the driver of acute respiratory distress syndrome (ARDS) and may lead to multi-organ dysfunction^22,23^.The activation of the complement system also can be found in patients infected with coronaviruses, such as MERs-COV, SARS-CoV-1 and SARS-CoV-2, which develop into ARDS^24^. When mice with complement C3 deficiency were infected with SARS-COV, the infiltration of neutrophils and inflammatory monocytes in the lungs was significantly reduced, and the levels of cytokines and chemokines in the lungs and serum were decreased, as well as the incidence of respiratory failure. This suggests that the activation of the complement component C3 may aggravate the disease of SARS-COV-related ARDS^23^. Young patients have fewer underlying diseases and are more immune-related compensated. This may be the reason why many indicators including D-dimer and procalcitonin may not change significantly even when they progress to severe COVID-19. However, young patients have more active immune function, which is why the serum complement C3 may be a potential indicator of the severity of COVID-19 patients.

This study possesses limitations aside from those inherent to all retrospective cohort studies such as their lack of causal inference. First, it is a single-center study featuring a limited number of cases. Second, the patient data were collected within 3 days after hospital admission, leading to missing data in a number of variables, which were imputed with a standard statistical approach.

In summary, this study conducted a comprehensive statistical analysis with a focus on non-linear relationships to identify risk factors and possible pathogenesis for the development of a severe disease course during COVID-19 infection in young patients. However, large-scale and multi-center analysis is needed to foster the obtained knowledge.

## Data Availability

The data can be obtained through the communication with corresponding author

## Contributors

Acquisition, analysis, or interpretation of data: WC, KX, RH, JL; Drafting of the manuscript: WC, RH, JL;

Statistical analysis: RH, JL;

Critical revision of the manuscript for important intellectual content: KH, KX; Obtained funding: WC, KX, RH;

Administrative, technical, or material support: WC, KX; Conception design: WC, KX, JL;

Supervision: WC, KX, JL.

## Ethics, consent and permissions

The study was approved by the Ethics Committee of Wuhan No.1 Hospital (No. 202008). All methods were carried out in accordance with relevant guidelines and regulations.

## Consent to publish

For this study, informed consent was obtained from all subjects or, if subjects are under 18, from a parent and/or legal guardian.

